# Job Satisfaction and Its Determinants Among Occupational Safety Experts Working in Turkiye’s Joint Health and Safety Units

**DOI:** 10.1101/2025.07.28.25331855

**Authors:** Ebru Kaya, Mahmut S. Yardim

**Affiliations:** Hacettepe University

**Keywords:** Job Satisfaction, Joint Health and Safety Units, Occupational Safety Experts, Turkiye

## Abstract

**Introduction:** This study aims to examine the job satisfaction of occupational safety experts working in Joint Health and Safety Units (JHSUs) in Turkiye, and to identify the associated factors, including perceived competence, workload, support, and training.

**Methods:** A cross-sectional descriptive study was conducted in September 2024 using an online questionnaire. A total of 102 occupational safety experts participated. The survey included sociodemographic questions, the Job Satisfaction Questionnaire, and the Warwick Edinburgh Mental Wellbeing Scale. Chi-square, post-hoc power, and regression analyses were used to explore associations between variables and job satisfaction.

**Results:** The findings indicated that mental wellbeing, perceived competence, managerial support, and the ability to fulfill job requirements were significantly associated with higher job satisfaction. Perceived adequacy of training and concerns about job security also played a role. Despite expectations, increasing years of experience and certification class did not correspond with increased job satisfaction, suggesting that experience alone may not enhance perceived competence. The overall sample was skewed toward more experienced and highly engaged professionals, with underrepresentation of C class certified experts.

**Conclusion:** The study provides insights into factors influencing job satisfaction among occupational safety experts and highlights the role of psychological and organizational dynamics beyond demographic characteristics. The findings underline the need for targeted strategies to improve training quality, support mechanisms, and working conditions. Furthermore, such research contributes to a broader understanding of how differing national OHS practices impact the job satisfaction and competencies of safety professionals.

## Introduction

Occupational health and safety (OHS) is defined as a discipline concerned with the prevention of work-related injuries and illnesses, as well as the protection and promotion of workers’ health [1]. OHS professionals play a critical role in the implementation and management of health and safety practices in workplaces. To ensure safe and healthy working conditions, employers may either assign internal OHS professionals or obtain support and consultation from independent experts or external OHS service providers [1].

According to the Regulation on Occupational Health and Safety Services in Türkiye, a Joint Health and Safety Unit (JHSU) must be authorized by the Ministry and equipped with the necessary personnel and equipment to deliver OHS services to workplaces that receive such services externally. Occupational safety experts (OSE) appointed in this manner often serve multiple workplaces and work part-time for each, depending on the number of employees and the hazard classification of the workplace.

Following the enactment of the Occupational Health and Safety Law in Türkiye in 2012, the assignment of OSEs became mandatory, leading to a significant increase in the demand for certified professionals. While there were 47,000 certified experts in 2014, this number rose to rapidly and as of June 2025, 152,257 OSEs were registered in the Ministry’s system [2, 3]. This increase contributed to the widespread establishment of JHSUs, which fulfilled the OHS professional needs of many companies. However, this form of employment has been found to be significantly more stressful than being a full-time staff member assigned to a single workplace [4].

Working conditions, environment, and hours have a significant impact on job satisfaction [5]. The existing literature on job satisfaction among OSEs—particularly those employed in JHSUs—is limited, especially in cross-national contexts, indicating a notable research gap.

### Occupational Safety Experts

Significant differences exist in the regulatory frameworks governing OHS professionals across countries. In nations such as Australia, Canada, New Zealand, and the United Kingdom, the regulations are flexible and principle-based, emphasizing general employer responsibilities rather than mandating specific OHS roles. In contrast, countries like France, Indonesia, Japan, South Korea, Vietnam, and Spain have detailed legal frameworks that specify the roles, educational requirements, and ongoing professional development of OHS professionals. Singapore and Thailand adopt a hybrid approach, offering both regulatory flexibility and specificity for certain roles. These variations underscore the diversity in how countries define and regulate the roles of OHS professionals [1].

In Turkiye, The Occupational Health and Safety Law, enacted in 2012, aimed to align with European Union directives and introduced a preventive and protective approach to OHS. The law encompasses essential elements for structured OHS practices in workplaces, including training, risk assessment, the identification of preventive measures, and workplace inspections [6]. Safety experts are defined as certified technical personnel from engineering or architecture backgrounds. Since 2015, OHS courses have been mandatory in relevant faculties to ensure basic professional knowledge.

According to the Turkish Regulation, the training program for safety experts includes both theoretical and practical components, comprising a total of 220 hours, including a 40-hour internship. Safety experts are classified into A, B, or C categories based on their competency to serve workplaces with different hazard levels. For each level, the required training must be completed, followed by a national examination to obtain the corresponding certificate.

The Regulation defines the responsibilities of safety experts across all certification levels, with the primary distinction being the hazard classification of the workplaces they are authorized to serve. Safety experts are expected to provide guidance to employers on work planning, the use of personal protective equipment, and compliance with safety regulations. They are also responsible for identifying preventive measures to reduce the recurrence of workplace accidents and occupational diseases, as well as investigating incidents and advising employers accordingly. Their duties further include participating in risk assessments, overseeing the implementation of safety measures, conducting workplace inspections, and coordinating emergency preparedness activities. Additionally, they are tasked with planning and delivering occupational health and safety training for employees. These responsibilities demonstrate the central role of safety experts in evaluating workplace health and safety conditions and supporting employers in meeting their legal obligations to maintain a safe working environment.

### Job Satisfaction and Mental Well-Being

Job satisfaction, defined as “a pleasurable or positive emotional state resulting from the appraisal of one’s job or job experiences” [7], is influenced by various factors including job characteristics such as skill variety, task identity, task significance, feedback, and autonomy, as well as stressors like workload, organizational constraints, interpersonal conflicts, and perceived injustice. The availability of resources, particularly social support, also plays a crucial role [8]. Among OSEs, intrinsic motivators such as helping others, professional prestige, autonomy, and opportunities to apply expertise are associated with higher satisfaction, whereas extrinsic challenges like inadequate salary, excessive workload, and unsupportive managerial behavior are common sources of dissatisfaction [9, 10]. Job satisfaction is inversely related to depression levels, with higher satisfaction linked to lower depression [11]. Demographic factors such as gender and marital status have shown some influence, with male and single experts reporting higher satisfaction compared to females and married individuals, while younger and less experienced professionals often report lower satisfaction [9,10].

Additionally, educational background and certification levels affect job satisfaction, with bachelor’s degree holders and B-class certified experts reporting higher satisfaction than their counterparts [10]. Mental well-being, closely linked to job satisfaction, reflects an individual’s emotional and cognitive evaluation of life and significantly affects their ability to cope with challenges, function effectively, and maintain social and professional engagement [12,13]. In line with the World Health Organization’s definition, well-being is a multidimensional construct shaped by social, economic, and environmental conditions, encompassing individuals’ capacity to live meaningful and fulfilling lives [14].

This study aims to investigate the relationships between the sociodemographic characteristics, work-related factors, OHS education background, competency assessments, mental well-being, and job satisfaction of OSEs employed by JHSUs. By identifying the factors influencing job satisfaction, the study seeks to offer recommendations for improving working conditions. Recognizing these factors is a critical step toward understanding the daily challenges faced by professionals and, ultimately, enhancing both the individual performance of OSEs and the quality of OHS conditions in workplaces receiving such services. The study received no external funding. All research expenses were covered by the authors.

### Methodology

This cross-sectional analytic study, employing descriptive and inferential statistical methods, was conducted among OSEs working in Joint Health and Safety Units (JHSU) in Turkiye. As of December, there are 2,485 JHSUs authorized by the Ministry of Labor and Social Security in Turkiye [3]. The study population consisted of 102 OSEs working in JHSUs.

The Job Satisfaction Scale was used to measure dependent variable, job satisfaction. This scale evaluates employees’ feelings, thoughts, and attitudes towards their work. It consists of four dimensions: Management Policies and Practices, Job Structure, Communication and Relations, and Physical Working Conditions, designed for white-collar samples. For OSEs working in Joint Health and Safety Units (JHSUs), the dimensions of Management Policies and Practices and Physical Working Conditions were excluded from the measurement of job satisfaction, as these professionals serve multiple workplaces, making evaluation of those dimensions impractical [15].

Mental well-being, assessed by the Warwick-Edinburgh Mental Well-Being Scale (WEMWBS) that includes positive items reflecting optimism, feeling useful, relaxation, interest in others, energy, problem-solving ability, clear thinking, good feelings, closeness to others, self-confidence, decisiveness, love, interest in new things, and cheerfulness, was treated as an independent variable influencing job satisfaction [16].

In addition to the main variables of interest, covariates comprised sociodemographic characteristics (age, gender, marital status) and occupational factors (e.g., previous OHS training and competency evaluations), which were considered potential confounders in the regression analysis.

An online survey was prepared and distributed to OSEs working in JHSUs. An email containing information about the study and the survey link was sent to 2,584 authorized JHSUs requesting that the email be forwarded to OSEs and that they complete the survey. Additionally, the survey was shared on different OSEs’ communication groups.

Job satisfaction scores were calculated by summing the Job Structure with Communication and Relations dimensions. Job Satisfaction level was divided into ‘Low’ and ‘High’ groups according to their median values. For comparisons between independent groups, Pearson’s chi-square test was used for categorical variables.

Statistical significance was set at p < 0.05. All statistical analyses were conducted using IBM SPSS Statistics version 23. In addition, a post-hoc power analysis was performed using GPower 3.1 software. Finally, a stepwise multiple regression analysis was carried out to identify the effects of independent variables on the dependent variable.

## Results

Of the respondents, 63.7% were male and 36.3% female. As shown in Table 1, 26.5% of participants were under 3 years old, 47.1% were between 36-45 years, and 26.5% were over 45. Among female participants, 51.4% were 35 or younger, compared to only 12.3% of male participants. Meanwhile, 55.4% of males were aged 36-45, and 32.3% were 46 or older. This age distribution difference between genders was statistically significant (p < 0.001).

**Tablo 1.**
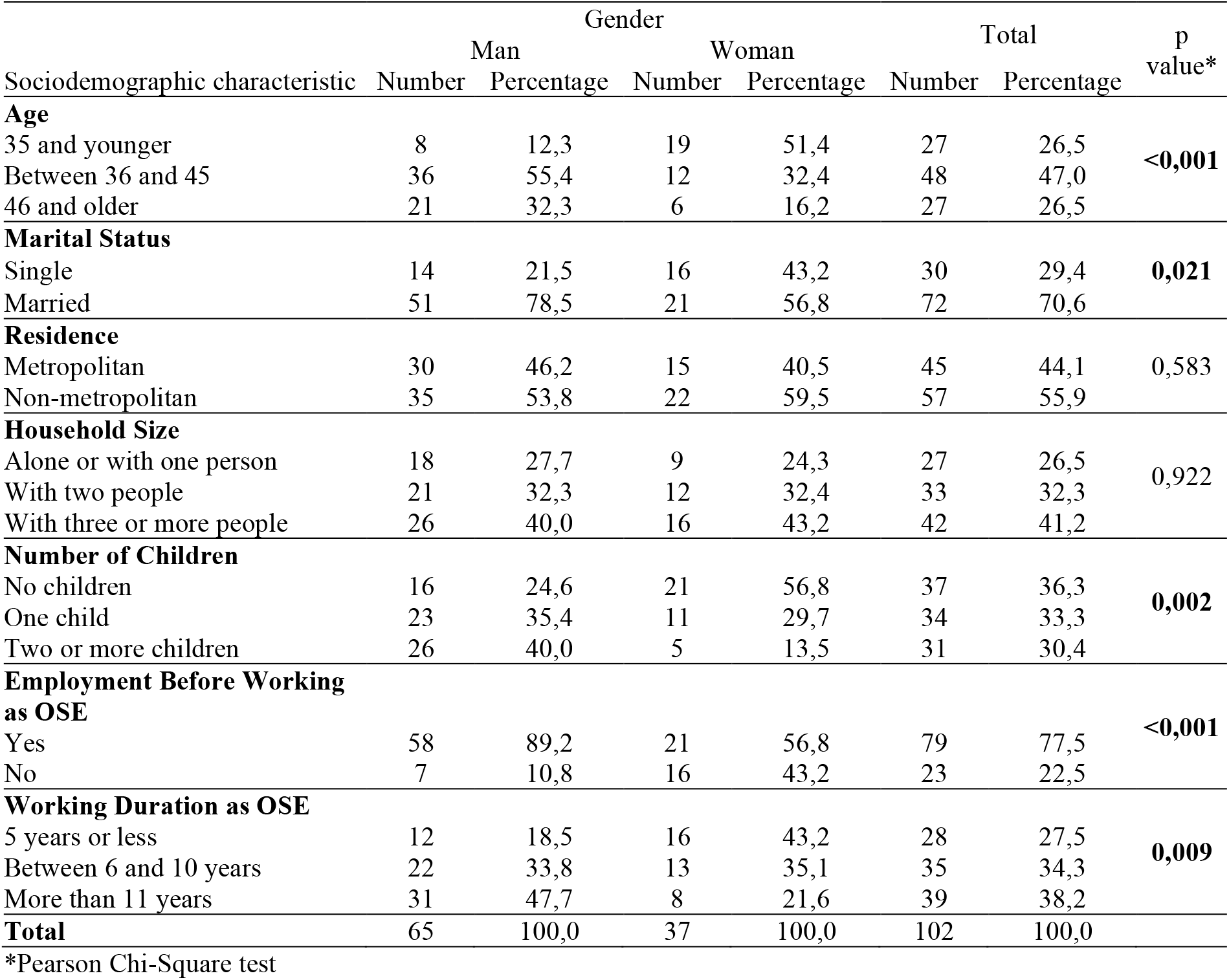
The distribution of sociodemographic characteristics of participants by gender.

70.6% of participants reported being married. The marriage rate was 78.5% for males and 56.8% for females, with a statistically significant difference in marital status by gender (p = 0.021).

36.3% of participants had no children. Among women, 56.8% had no children and 29.7% had one child. Among men, 24.6% had no children, and 40% had two or more children. The difference in number of children between genders was statistically significant (p = 0.002).

Participants were asked if they had worked in another profession before becoming OSEs. According to Table 1, 77.5% stated that occupational safety was not their first profession. Among women, 56.8% indicated this, while the rate was 89.2% for men (p < 0.001).

Regarding length of employment as an occupational safety expert, 27.5% had been working for 5 years or less, 34.3% for 6-10 years, and 38.2% for more than 11 years. Among women, 43.2% had worked 5 years or less, compared to only 18.5% of men (p = 0.009).

After completing the survey, participants were asked to provide opinions, suggestions, and comments through an open-ended question. Responses were categorized by the researcher, and participants were allowed to mention multiple issues. The most frequently raised topic was salary (31.0%), followed by enforcement authority (29.6%) (see Table 2).

**Tablo 2.**
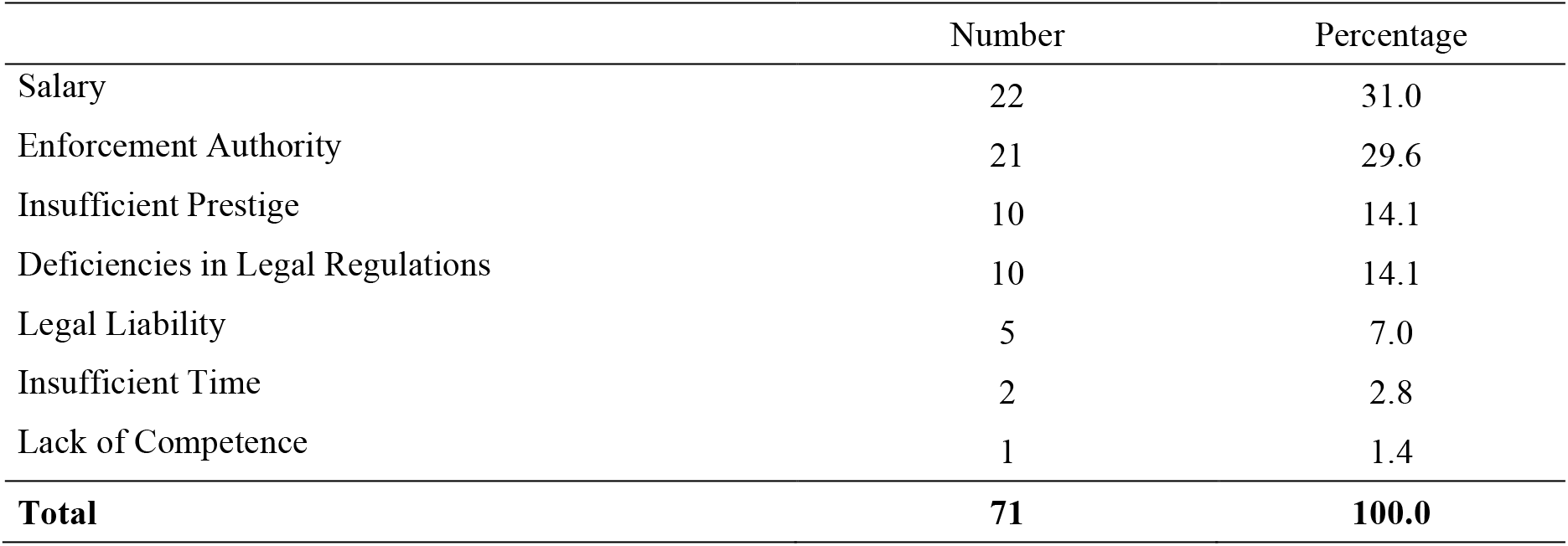
Distribution of Participants’ Opinions and Suggestions by Topic.

Chi-square tests were conducted to examine the relationships between various variables and job satisfaction. Table 3 presents the statistical analysis results for all variables evaluated for their impact on job satisfaction. Due to the sample size of 102 in this study, many variables that appeared to be related in the cross-tabulations were not found to have statistically significant effects. To better understand the influence of these variables on the target variable, the tables also include Cramer’s V values indicating effect size, along with the recommended minimum sample size based on these effect sizes.

**Tablo 3.**
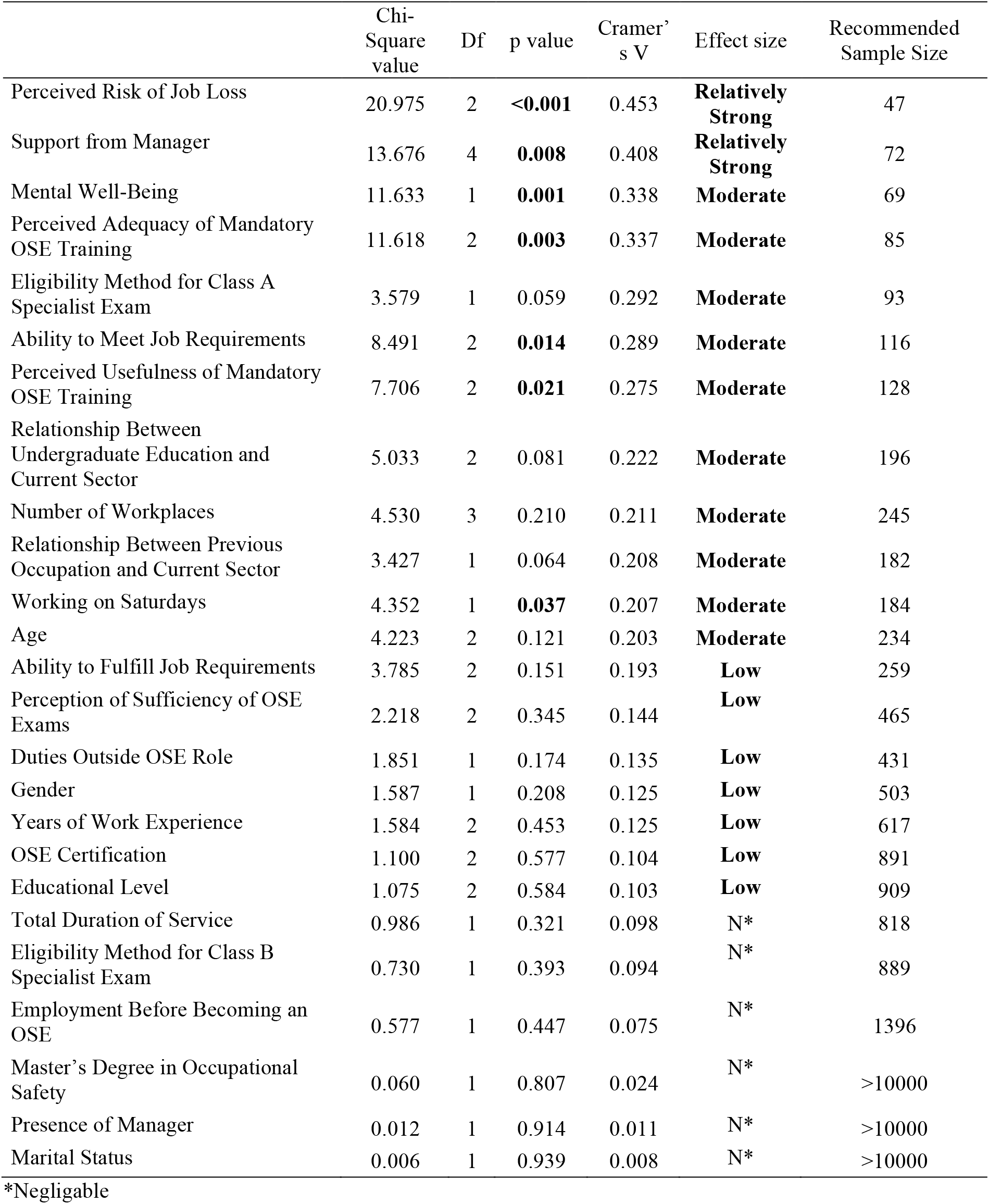
Chi-Square and Post-Hoc Power Analysis Results for Variables Potentially Associated with Job Satisfaction.

Cramer’s V values were interpreted as follows: values between 0 and 0.10 indicate a negligible effect; 0.10 to 0.20 indicate a weak effect; 0.20 to 0.40 indicate a moderate effect; 0.40 to 0.60 indicate a relatively strong effect; 0.60 to 0.80 indicate a strong effect; and 0.80 to 1.00 indicate a very strong effect [17].

According to Table 3, variables with a statistically significant impact on job satisfaction include working on Saturdays, perceived usefulness of OSE training, perceived adequacy of OSE training, and ability to meet job requirements, all of which have moderate effect sizes. Additionally, perceived risk of job loss and managerial support have relatively strong effect sizes.

Furthermore, for variables with moderate effect sizes—such as age, eligibility method for class A specialist exam, number of workplaces, relationship between previous occupation and current sector, and relationship between undergraduate education and current sector —statistically significant relationships may be detected if the sample size is increased to the levels recommended in the table.

Based on the results of the bivariate analyses, variables with statistically significant associations (p < 0.05) and at least moderate effect sizes (Cramer’s V ≥ 0.25) were selected as candidates for inclusion in the stepwise multiple regression model. These included perceived risk of job loss, support from manager, mental well-being, perceived adequacy and usefulness of mandatory OSE training, and the ability to meet job requirements. This selection strategy balances statistical rigor with parsimony, considering the study’s sample size (n = 102) and the potential for overfitting. Variables with weak or non-significant associations (e.g., p > 0.10 and Cramer’s V < 0.25) were excluded to maintain model stability and interpretability.

As a result of the regression analysis conducted with the variables describing Job Satisfaction, the adjusted R-squared value—indicating the predictive power of the selected variables—was calculated as 0.458.

According to the results, a one-unit increase in the perceived risk of job loss in the next 12 months is associated with a 1.58-point decrease in Job Satisfaction. A one-point increase in the perceived adequacy of the mandatory occupational safety training taken prior to the certification exam leads to a 2.26-point increase in Job Satisfaction. Similarly, a one-unit increase in the variable representing the ability to meet the job’s requirements results in a 2.18-point increase in Job Satisfaction. Lastly, a one-unit increase in Mental Well-Being Score is associated with a 0.27-point increase in Job Satisfaction.

**Tablo.** Hata! Belgede belirtilen stilde metne rastlanmadı.. Job Satisfaction Regression Analysis.

| Descriptive Variables for Job Satisfaction | Unstandardized Coefficients |  | Standardized Coefficients | p value |
| --- | --- | --- | --- | --- |
|  | B | Standard Error | Beta |  |
| Constant | 29.76 | 3.87 |  | <0.01 |
| Perceived Risk of Job Loss Within the Next 12 Months | -1.58 | 0.43 | -0.29 | <0.01 |
| Perceived Adequacy of Mandatory OSE Training Prior to Certification Exam | 2.26 | 0.83 | 0.22 | 0.01 |
| Ability to Meet Job Requirements | 2.18 | 0.94 | 0.20 | 0.02 |
| Mental Well-Being Score | 0.27 | 0.08 | 0.31 | <0.01 |

Adjusted R-Square = 0.458 Model F value = 22.32; p value < 0.001

## Discussion

Among participants, 63.7% were male and 36.3% were female. Compared to previous studies, this sample contains a higher proportion of male respondents. According to Turkish Statistical Institute data, women constitute 34% of the employed population [18]. A 2020 study on emotional well-being among OSEs reported that 41% of participants were female [19], while a 2022 study on job satisfaction levels found the figure to be 36% [9]. The gender distribution in this study appears to align with prior research, suggesting that the sample adequately reflects the gender distribution of the occupational safety expert population. The predominance of male participants may reflect the gender imbalance in the profession.

Additionally, 51.4% of female participants were under the age of 35, whereas this figure was only 12.3% for males. In terms of work experience, 81.5% of male participants had over five years of experience, compared to 56.2% of female participants. This may suggest that female experts tend to leave the workforce after age 35.

Supporting this observation, 56.8% of female participants were married, compared to 78.5% of males. These data suggest that women may leave the profession after marriage. According to Turkish Statistical Institute, the average career duration is 20.2 years for women and 39.2 years for men [18]. These findings are consistent with broader national trends indicating that male professionals pursue greater career continuity.

The occupational safety expert certification is available to individuals who have graduated from engineering, architecture, and fundamental science disciplines. The majority of professionals currently working in this field did not initially plan to pursue this career during their university education. In fact, 77.5% of participants indicated that occupational safety was not their first profession. Following the implementation of mandatory employment of OSEs in all workplaces, many individuals obtained certification and transitioned into this role from other fields. This indicates that university-level education alone may not sufficiently prepare individuals for this profession, necessitating additional training and certification. In this regard, occupational safety training programs and certification exams serve as important tools for enhancing professional competence.

However, 65.7% of participants reported that they found the occupational safety training they received to be inadequate, while 15.7% were undecided. Only 18.6% considered the training sufficient. The transition to remote education and the removal of refresher training requirements may have contributed to this perception of inadequacy.

Approximately 58% of participants responded to the open-ended question soliciting their views and suggestions. That a majority of OSEs took additional time to respond—often with lengthy and thoughtful answers—after completing the structured part of the survey indicates both a strong awareness of issues within the profession and a willingness to contribute to potential solutions. Additionally, responses to this open-ended question could be grouped into seven thematic areas, indicating consistency among respondents in identifying and articulating common challenges.

The most frequently mentioned concern was inadequate remuneration. Respondents noted that the lack of a standardized minimum wage policy results in lower compensation for OSEs employed through Joint Health and Safety Units (JHSUs). Several participants stated that problems arise from the structure in which employers pay for occupational safety services through JHSUs, rather than directly to the experts. This commercial relationship encourages price-based competition among JHSUs, which in turn lowers both the financial compensation for experts and the perceived value of the service. Respondents also reported practices such as partial off-the-books payments and irregular salary disbursements. A study on workplace physicians employed through JHSUs found that 63.9% of participants received only part of their agreed-upon salary [20]. Similarly, other field studies have shown that the lack of a minimum fee for services provided through JHSUs has led to an environment characterized by cheap and undervalued safety services [21].

Many participants felt that the competitive pricing strategies employed by JHSUs negatively affect the professional dignity of the occupational safety expert role. Respondents expressed the belief that they are not respected professionally, and that employers only obtain occupational safety services due to legal obligations. Another factor contributing to the perceived lack of professional respect is that many experts did not voluntarily choose this career path. In a qualitative study involving face-to-face interviews, 57% of OSEs stated that they entered the profession because they were unable to find jobs in their trained fields or failed to be appointed to public sector positions in those areas [22].

The second most frequently mentioned issue was the lack of enforcement power over employers. OSEs reported difficulties in ensuring that the hazards they identify in the workplace are adequately addressed by employers.

Those employed through JHSUs noted that when they attempt to document safety deficiencies in the logbook or report them to the Ministry, employers often respond by requesting a change in the assigned safety expert through the JHSU. Additionally, JHSU-employed safety experts do not have the right to choose the workplaces they serve. These conditions undermine the professional autonomy of OSEs.

Participants also expressed dissatisfaction with the inadequacy of legal regulations and the lack of enforcement, even when regulations are in place. The insufficiency of ministry-led inspections was seen as a factor that further weakens the influence and authority of OSEs. In a 2018 study, gaps, deficiencies, and delays in occupational safety legislation were also identified as major concerns [23].

Another serious issue raised by participants was the legal liability faced by OSEs in the event of a workplace accident. When considered alongside their lack of perceived autonomy and limited enforcement power, the fact that experts can be held legally responsible—even receiving criminal sentences—highlights the precarious nature of their professional responsibilities.

Many experts also commented on the limited time they have to fulfill their responsibilities, especially those employed through JHSUs who must serve multiple workplaces. In cases where more than one site must be visited in a single day, time spent commuting is not included in official working hours, further limiting their capacity to perform their duties effectively. OSEs think limiting service duration alone is insufficient, and also recommended restricting the number of workplaces served by a single expert [4]. Such limitations could not only help experts fulfill their legal obligations more thoroughly but also provide them with the opportunity to engage in professional development activities, thereby improving their expertise and overall competence.

The R-square value in the model developed for Job Satisfaction was 0.458, indicating that the independent variables had a substantial explanatory power. Furthermore, the standardized beta coefficients of the statistically significant predictors suggest notable individual effects of these variables.

A statistically significant relationship was observed between job satisfaction and the perceived risk of job loss within the next 12 months. Job insecurity was found to have a statistically significant negative impact on job satisfaction [24, 25].

A statistically significant relationship was also found between the perceived adequacy of mandatory OSE training prior to certification exam and job satisfaction. Participants who found the training sufficient reported higher job satisfaction. Feeling competent in their specific occupational responsibilities was associated with increased job satisfaction [26]. The ability to meet job requirements and the sense of being useful were also positively related to job satisfaction.

Mental well-being and gender were also found to significantly affect job satisfaction suggesting that individuals with higher mental well-being are more likely to find meaning in their work.

No statistically significant relationship was found between certification levels (C, B, A class) and job satisfaction. This contradicts with the findings reported that B-class experts had the highest job satisfaction and A-class experts the lowest [10]. The discrepancy may be due to temporal factors: in 2018, only five years had passed since the enactment of the Occupational Health and Safety Law, and many A-class experts had obtained their certifications via direct examination rather than field experience. Therefore, although they worked in high-risk workplaces, their perceived competence may not have matched the job’s demands, which could have negatively influenced satisfaction. In our study, the lack of differentiation in job satisfaction across certification levels may stem from the similarity of job content and the absence of corresponding increases in professional autonomy or competence.

### Limitations

The inability to distribute the survey via the Ministry’s national online platform limited participant recruitment, resulting in a sample of 102 OSEs. This restricts the generalizability of the findings and the ability to perform advanced statistical analyses. Nevertheless, comparative analyses were conducted to provide insights that may inform future studies, though causal interpretations remain limited.

The reliance on self-reported data introduces subjectivity, and the sample’s relatively small size necessitates caution in generalizing results to all JHSU-employed OSEs. Additionally, the online distribution through professional networks may have biased participation toward individuals with higher engagement and satisfaction levels.

There was also an imbalance in certification levels: only 18.6% of respondents were C-class experts, despite this group constituting 61% of all registered OSEs [3]. The underrepresentation of this relatively new professional group may have influenced the results.

Finally, responses concerning training adequacy and exam effectiveness reflected personal perceptions rather than objective assessments, and should be interpreted accordingly.

## Conclusion and Implications

Due to the relatively recent emergence of the occupational safety expert profession, the lack of traditional master-apprentice learning experiences, insufficient remuneration, absence of enforcement authority, low professional prestige, and the associated legal liabilities, it is expected that job satisfaction in this field is relatively low. Only 8.0% of participants reported working as OSEs prior to the enactment of the Occupational Health and Safety Law. Despite these challenging conditions, OSEs view their profession as meaningful, consider the training they receive effective, and find the examinations beneficial. Their dedicated efforts play a significant role in reducing serious workplace accidents in the country.

Further studies should explore whether there is a relationship between job satisfaction and professional competence. Given that increased competence in their profession likely leads to greater job satisfaction and enhanced contributions to workplace safety, promoting competency development may positively impact the occupational safety culture in Turkiye. No relationship was observed between job satisfaction and holding a master’s degree in occupational safety. Research is needed to investigate the role of postgraduate education in enhancing professional competence.

The quality and effectiveness of training received by OSEs prior to certification examinations should be regularly monitored. Additionally, supplementary training opportunities for these experts should be increased.

In the study, 59.8% of participants expressed dissatisfaction with the adequacy of the occupational safety expert (OSE) exams. The validity and reliability of certification exams in measuring competency should be evaluated to ensure their effectiveness.

No significant relationship was found between occupational safety expert certification levels or work experience in years and job satisfaction. Although prior studies have shown that increased competence correlates with higher job satisfaction, this study did not observe such differences specific to the OSE profession.

This and similar studies can contribute to understanding how national differences in occupational health and safety practices influence the job satisfaction and competencies of occupational safety specialists, offering useful insights for developing context-specific strategies and improving international policy alignment.

## Data Availability

All data produced in the present study are available upon reasonable request to the authors

## References

1. International Labour Organization. Occupational safety and health professionals at the workplace level: A review of qualification systems and regulatory approaches in selected countries. Geneva: International Labour Organization; 2022.

2. Çalışma ve Sosyal Güvenlik Bakanlığı. İş güvenliği uzman sayısının 165 bini aştığını açıkladı. [Internet]. 2020 [cited 2024 Dec 12]. Available from: https://www.csgb.gov.tr/isggm/haberler/is-guvenligi-uzman-sayisinin-165-bini-astigini-acikladi/

3. T.C. Çalışma ve Sosyal Güvenlik Bakanlığı. İSG-Katip. [Internet]. [cited 2025 Jul 6]. Available from: https://isgkatip.csgb.gov.tr/anasayfa

4. Karakaya T. İş Güvenliği Uzmanlarının Çalışma Yaşamı Özellikleri İş Stresi ve İş Güvencesizliğinin Değerlendirilmesi [master’s thesis]. Ankara: Hacettepe Üniversitesi; 2018.

5. Aslan M, Uysal Ş, Oğuzhan Y. Uzaktan çalışma, iş yaşam dengesi, iş doyumu ve yaşam doyumu arasındaki ilişkilerin teorik perspektiften incelenmesi. 5th International Paris Conference on Social Sciences; 2021.

6. Alpagut G. 6331 Sayılı İş Sağlığı ve Güvenliği Kanununun Genel Esasları. İÜHFM. 2014;72(2):31–46.

7. Locke EA. The nature and causes of job satisfaction. In: Dunnette MD, editor. Handbook of industrial and organizational psychology. Chicago: Rand McNally; 1976. p. 360–580.

8. Meier LL, Spector PE. Job satisfaction. In: Wiley Encyclopedia of Management. Vol. 5. 2015.

9. Toyoğlu AE. İş Güvenliği Uzmanlarının İş Tatmin Düzeyini Etkileyen Bireysel Faktörlerin İncelenmesi [master’s thesis]. 2022.

10. Cerev G. İş güvenliği uzmanlarının genel, içsel ve dışsal iş tatmin düzeylerinin incelenmesi üzerine bir araştırma. Yönetim Bilimleri Dergisi. 2018;15(30):91–112.

11. Atalı G, Ağar A, Acar MN. İSG profesyonellerinin depresyon düzeyleri ve iş tatmin düzeyleri arasındaki ilişkinin incelenmesi. Sosyal Güvence Dergisi. 2023;23:969–87.

12. Mental Health Foundation. What is mental health? [Internet]. 2008 [cited 2025 Jan 15]. Available from: https://www.mentalhealth.org.uk/explore-mental-health

13. Dinesh B, Till A, Sartorius N. What is mental health? Int J Soc Psychiatry. 2013;59(1):3–4.

14. World Health Organization. Health Promotion Glossary of Terms. Geneva: WHO; 2021.

15. Ünsal P, Türetgen İÖ. Bir iş doyumu ölçeği geliştirme çalışması. Yönetim. 2005;16(50):43–55.

16. Keldal G. Warwick-Edinburgh Mental İyi Oluş Ölçeği’nin Türkçe formu. J Happiness Well-Being. 2015;3(1):103–15.

17. Lee DK. Alternatives to P value: confidence interval and effect size. Korean J Anesthesiol. 2016;69(6):555– 62. doi:10.4097/kjae.2016.69.6.555

18. Türkiye İstatistik Kurumu (TÜİK). [Internet]. 2025 [cited 2025 Jan 30]. Available from: https://data.tuik.gov.tr

19. Aytaç S, Engin T, İmanlı E. İş güvenliği uzmanlarının işe ilişkin duygusal iyi oluş hali mutluluk ve yaşam tatmini ilişkisi. J Yasar Univ. 2020;15:746–58.

20. Türkiye Tabipler Birliği. İşçi sağlığı ve işyeri hekimliği. Ortak Sağlık ve Güvenlik Biriminde çalışan hekimlerin çalışma koşulları ve yaşadıkları sorunlar. Mesleki Sağlık ve Güvenlik Dergisi. 2018;Nisan– Eylül.

21. Taşkıran G. Güvencesiz iş güvenliği uzmanları, piyasalaşan iş güvenliği: bir alan araştırması. Çalışma ve Toplum. 2016;4.

22. Aca Z, Göze F. OSGB sistemiyle çalışan iş güvenliği uzmanlarının iş güvenliği uzmanlığına ilişkin görüşleri. Çalışma ve Toplum. 2020;4.

23. Takaoğlu Başkan Z, Kaya Çelenk E, İri Ölmezoğlu Nİ. İş güvenliği uzmanlarının yaşadığı sorunlar. Gümüşhane Univ Sağlık Bilim Derg. 2018;7:152–67.

24. Kaya İ, Artz B. The impact of job security on job satisfaction in economic contractions versus expansions. Appl Econ. 2015;47(24):2471–90.

25. Zheng X, Diaz I, Tang N. Job insecurity and job satisfaction. Career Dev Int. 2014;19(3):293–312.

26. Dharmanegara I, Sitiari N, Wirayudha I. Job competency and work environment: the effect on job satisfaction and job performance among SMEs workers. IOSR J Bus Manag. 2016;18(1):19–26.

